# Therapeutic potential of IL6R blockade for the treatment of sepsis and sepsis-related death: Findings from a Mendelian randomisation study

**DOI:** 10.1101/2022.07.14.22277638

**Authors:** Fergus Hamilton, Matt Thomas, David Arnold, Tom Palmer, Ed Moran, Alexander J Mentzer, Nick Maskell, Kenneth Baillie, Charlotte Summers, Aroon Hingorani, Alasdair MacGowan, Golam M Khandakar, Ruth Mitchell, George Davey Smith, Peter Ghazal, Nicholas J Timpson

**Affiliations:** MRC Integrative Epidemiology Unit, University of Bristol; Infection Science, North Bristol NHS Trust; Intensive Care Unit, North Bristol NHS Trust; Academic Respiratory Unit, University of Bristol; Wellcome Centre For Human Genetics, University of Oxford; Roslin Institute, University of Edinburgh; Department of Medicine, University of Cambridge; UCL Institute for Cardiovascular Science, University College London; UCL BHF Research Accelerator, University College London; Health Data Research UK; Project Sepsis, Cardiff University

## Abstract

**Introduction:** Sepsis is characterised by dysregulated, life-threatening immune responses, which are thought to be driven by cytokines such as interleukin-6 (IL-6). Genetic variants in *IL6R* known to downregulate IL-6 signalling are associated with improved COVID-19 outcomes, a finding later confirmed in randomised trials of IL-6 receptor antagonists (IL6RA). We hypothesised that blockade of IL6R could also improve outcomes in sepsis.

**Methods:** We performed a Mendelian randomisation analysis using single nucleotide polymorphisms (SNPs) in and near *IL6R* to evaluate the likely causal effects of IL6R blockade on sepsis, sepsis severity, other infections, and COVID-19. We weighted SNPs by their effect on CRP and combined results across them in inverse variance weighted meta-analysis, proxying the effect of IL6RA. Our outcomes were measured in UK Biobank, FinnGen, the COVID-19 Host Genetics Initiative (HGI), and the GenOSept and GainS consortium. We performed several sensitivity analyses to test assumptions of our methods, including utilising variants around *CRP* in a similar analysis.

**Results:** In the UK Biobank cohort (N=485,825, including 11,643 with sepsis), IL6R blockade was associated with a decreased risk of sepsis (OR=0.80; 95% CI 0.66-0.96, per unit of natural log transformed CRP decrease). The size of this effect increased with severity, with larger effects on 28-day sepsis mortality (OR=0.74; 95% CI 0.38-0.70); critical care admission with sepsis (OR=0.48, 95% CI 0.30-0.78) and critical care death with sepsis (OR=0.37, 95% CI 0.14 - 0.98) Similar associations were seen with severe respiratory infection: OR for pneumonia in critical care 0.69 (95% CI 0.49 - 0.97) and for sepsis survival in critical care (OR=0.22; 95% CI 0.04- 1.31) in the GainS and GenOSept consortium. We also confirm the previously reported protective effect of IL6R blockade on severe COVID-19 (OR=0.69, 95% 0.57 - 0.84) in the COVID-19 HGI, which was of similar magnitude to that seen in sepsis. Sensitivity analyses did not alter our primary results.

**Conclusions:** IL6R blockade is causally associated with reduced incidence of sepsis, sepsis related critical care admission, and sepsis related mortality. These effects are comparable in size to the effect seen in severe COVID-19, where IL-6 receptor antagonists were shown to improve survival. This data suggests a randomised trial of IL-6 receptor antagonists in sepsis should be considered.

## Introduction

Sepsis is a complex physiological and metabolic response to infection, characterised by elevated levels of cytokines and organ dysfunction.^1^ Our current best treatments remain antimicrobial therapy and organ support, with no licenced treatments outside these interventions.^2^

Interleukin-6 (IL-6) is a critical cytokine involved in the innate immune response in sepsis and other severe infections and contributes in conjunction with other pathophysiological processes to adverse outcomes.^3–6^ The modulation of IL-6 dynamics and its multiple different signalling pathways represents a potentially exciting therapeutic opportunity for severe infection given the key role for this cytokine and associated signalling.^3,6,7^ The inflammatory role of IL-6 is mediated not by the classical or trans-presentation modes of action through a membrane bound receptor (IL6R), but through a trans-signalling mechanism where IL-6 binds a soluble form of the receptor that subsequently interacts with membrane bound signalling molecule, gp130, on cells.^7,8^

Inhibition of both membrane and soluble IL6R using monoclonal antibodies such as tocilizumab or sarilumab (collectively known as IL-6 receptor antagonists, IL6RAs) have been successfully trialled in critically ill patients with COVID-19 and are now considered a standard treatment.^9–11^ These drugs have the capability of attenuating all forms of IL-6 signalling, producing reductions in C-reactive protein (CRP) and other downstream inflammatory markers.^11–13^ Furthermore, this beneficial effect of IL6R blockade in COVID-19 was anticipated in a causal framework analysis using genetic data.^14,15^ Carriers of certain single nucleotide polymorphisms (SNPs) around and in the *IL6R* gene (the target for IL6RAs) that phenocopy IL6RA function have a reduced risk of becoming critically ill with COVID-19.^14,15^

The same *IL6R* variants have been used in Mendelian randomisation (MR) studies as instruments for alterations in IL6R function and signalling, thus providing a functional proxy of IL6RA therapy.^14,16–21^ In particular, a previous MR analysis suggested that reduced IL6R signalling could reduce inflammation and risk of coronary heart disease,^12^ leading to clinical trials of monoclonal antibodies targeting either IL6R or IL-6 for coronary disease prevention.^17^

We hypothesised that there may be a role for IL6R blockade in sepsis given the similarities between bacterial sepsis and critical illness in COVID-19,^22^. To test this, we undertook a two sample Mendelian randomisation study to assess the potential impact of IL6R blockade on sepsis, COVID-19, as well as risk of infection in the absence of sepsis.

## Methods

### Study design

In this study, we aimed to perform a Two Sample Mendelian randomisation study in order to proxy the effect of IL6R blockade on sepsis and other infections. **Figure 1** shows the overall study design, comparing our analysis with a randomised controlled trial of IL-6 receptor antagonist therapy.

**Figure 1:**
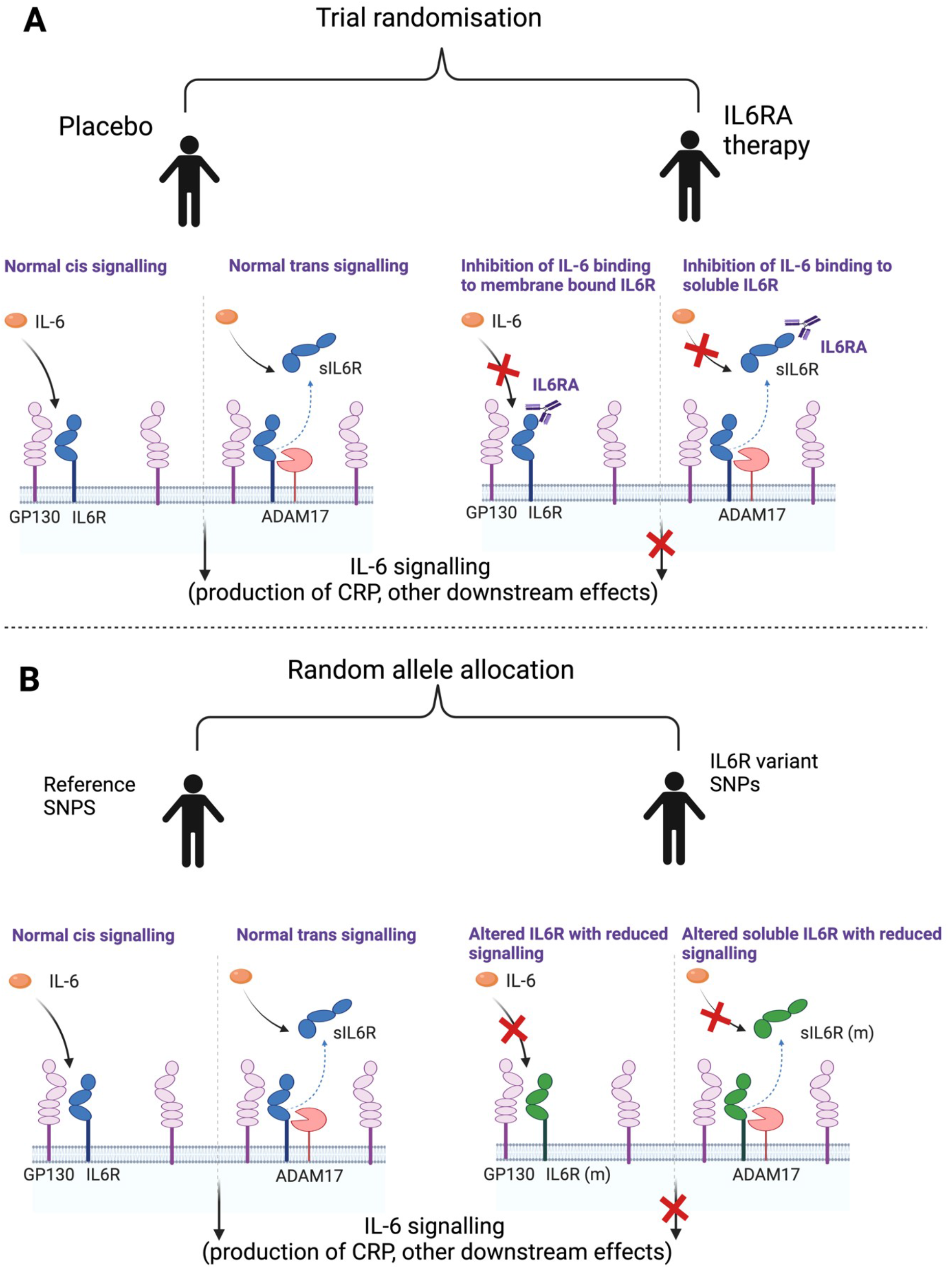
Section A represents a randomised trial of IL6RA therapy with both normal IL6 signalling and the effect of intervention. In Section B, represents the use of SNPs in *IL6R* to act as a proxy for intervention. We identify variants within *IL6R* associated with reduced CRP as a marker of functional IL6R blockade to aid the interpretation of Mendelian randomisation analysis. As these SNPs are within IL6R, we make the assumption that these SNPs have an effect through alteration of IL6R (either through modification of the protein itself or altering quantity of the protein). Abbreviations: IL6R: interleukin-6 receptor, sIL6R: soluble interleukin-6 receptor, IL6RA: interleukin-6 receptor antagonist, IL6R (m): modified interlekin-6 receptor, GP130: Glycoprotein 130: ADAM17: A disintegrin and metalloprotease 17

### Included populations

For our main outcomes, we used UK Biobank, a large UK adult volunteer cohort described in detail elsewhere.^23^ UK Biobank has linked genetic and physical data with direct links to national health care datasets. For secondary outcomes we utilised FinnGen (Round 6), a large prospective cohort study in Finland, linked to electronic health record data.^24^ For measurement of COVID-19 outcomes, we included data from the COVID-HGI (Round 7), a large meta-analysis of multiple studies including participants with COVID-19.^25^

Finally, for additional data on sepsis survival, we utilised summary statistics from a previous GWAS on survival from sepsis, which included data from the GaINS and GenOSept consortium.^26^ Full details of each cohort, inclusion criteria, and genetic quality control are available in the **Supplementary Methods**.

### Definition of outcomes

Our primary outcomes were the incidence of sepsis, sepsis requiring critical care utilisation, and 30-day mortality after an episode of sepsis or sepsis requiring critical care admission, measured in UK Biobank.

For secondary outcomes, we included a) a set of 9 other common infections that present to primary or/secondary health care (**Supplementary Table S2**) and c) COVID-19 infection, as a comparator.

Admissions with sepsis were identified in UK Biobank in ICD-coded linked secondary care data. ICD-10 codes A02, A39, A40 and A41 were used to identify sepsis, in line with recent literature.^27^ Cases were included if the code was in the primary or secondary diagnostic position in Hospital Episode Statistics (HES) data (or similar datasets in the devolved nations), provided by UK Biobank. We did not include self-reported cases, or cases only occurring in primary care. To ensure no contamination with COVID-19 related codes, we excluded codes for sepsis that occurred after 1^st^ February 2020.

Other infections were defined similarly and included by the presence of ICD-10 codes, derived by two authors (FH, DA), with a code list available in the **Supplementary Table S2**. Codes were derived from recent publications but altered to reflect 3-digit ICD-10 coding available in HES. Controls were defined by the absence of the ICD code.

For the definition of critical illness related to infection, we utilised critical care admission data provided in HES. We considered any critical care episode during the index infection admission as a critical care admission related to that infection. Controls were defined as all other participants of UK Biobank. As HES data is only available for English participants of UK Biobank, we excluded all participants who were not recruited at a recruitment centre in England for the critical care portion of the study.

We generated 28-day survival outcomes for sepsis (with or without critical care admission), and Lower Respiratory Tract Infection (LRTI) or pneumonia (only in those admitted to critical care). These two infections were chosen as there were not enough cases (<350) to perform GWAS for other infections. Dates of admission to hospital were extracted from the HES data, and matched to national registry death data, supplied by UK Biobank.

In order to attempt to avoid issues of bias relating to the structure of the data included in primary analysis (in particular collider bias where selection on case status can induce associations between traits related to case status), we included all participants in our analysis of sepsis risk. For our analysis, we chose to use the whole UK Biobank population as a control for each outcome, even for the critical care related outcomes (e.g. we compared critically ill patients with sepsis vs the whole population, rather than critically ill patients with sepsis vs all patients with sepsis).

### GWAS of infection outcomes

To generate summary statistics for downstream MR, we performed a case-control GWAS on each infection outcome using regenie v2.2.4, on all UK Biobank participants of European ancestry (see critical care exclusions above).^28^ We used the in-house MRC-IEU GWAS pipeline (version 2) to quality control our data.^29^ Full details of the this are published elsewhere,^29^ with further details in the **Supplementary Methods**.

### Mendelian randomisation - Definition of instruments

Our *IL6R* instruments were selected based on a recent Mendelian randomisation study which performed a meta-analysis of a high sensitivity CRP Genome wide association study (GWAS) of 522 681 European individuals from the Cohorts for Heart and Aging Research in Genomic Epidemiology (CHARGE) Consortium and UK Biobank.^30^

Conceptually our genetic instrument is similar to the action of anti-IL6R monoclonal antibodies (e.g. tocilizumab) that lead to complete inhibition of IL6R signalling by blocking both IL6 classical and trans signalling.^17–19,31,32^

Specifically, we aimed to assess the effect of decreased activity of the IL6/IL6R pathway, modelled using independent (r^2^ <0.1) variants within 300kb of *IL6R* in order to proxy IL6RA effect. We call this our *cis*IL6R instrument. We weighted this variant on the effect on high sensitivity CRP, as justified in the **Supplementary Methods**, and in line with previous analyses. ^17–19^ For simplicity we henceforth refer to this exposure as “IL6R blockade”.

It is recognised that this is an oversimplification of the IL6 pathway, with evidence that effects on health outcomes are mediated by classical and trans signalling in differing ways, and that the effect of our instrument may not act in the same way as IL6RAs.^21^ In our sensitivity analyses (below), we explore other ways of defining our exposure, including alternative weighting strategies. In total, 26 variants were included. The minimum F-statistic of an included variant was 31.1.^33^ All SNPs were available in the GWAS performed above. The included SNP list is available in **Supplementary Table S1**.

### Statistical analysis

For our main analysis, we identified and extracted SNPs (or proxies, for secondary cohorts) from our outcome GWAS, and performed two sample Mendelian randomisation using harmonised SNPs on each outcome in term. MR estimates from each SNP were generated and then meta-analysed by inverse variance weighting.^32^ Analyses were performed using the *TwoSampleMR* package (version 0.5.6) in R (version 4.1.3).^34^

For the UK Biobank and FinnGen outcomes we performed fixed effects meta-analysis across each specific infection, to generate summary estimates for each condition, using the R package *meta*.

### cisCRP variants

Our primary instrument includes only variants around *IL6R*, weighted by their effect on CRP, as a read-out of IL-6 function. However, this does not imply that CRP itself is causal. In order to understand whether CRP might be the causal target, we undertook a subsequent analysis using four well-understood *cis*CRP related variants from a recent study (**Supplementary Table 1**).^35^ These variants are highly likely to alter CRP through a pathway independent of IL-6 downregulation, and therefore evidence of any MR effect would support CRP as a potential causal mechanism, and also provide additional genetic support for targeting of this pathway. These variants were weighted using the same UK Biobank-CHARGE meta-analysis of high sensitivity CRP.^36^

**Table 1:** Inverse variance weighted meta-analysis of MR estimates of IL6R blockade for UK Biobank and COVID-19 HGI outcomes.

|  | OR (95% CI) | P value | Cases / controls |
| --- | --- | --- | --- |
| Sepsis |  |  |  |
| Sepsis (all admissions) | 0.8 (0.66-0.96) | 0.019 | 11,643 / 474,841 |
| Sepsis (28 day death) | 0.74 (0.47-1.15) | 0.180 | 1,896 / 484,588 |
| Sepsis (critical care) | 0.48 (0.3-0.78) | 0.003 | 1,380 / 429,985 |
| Sepsis (28 day death in critical care) | 0.37 (0.14-0.98) | 0.046 | 347 / 431,018 |
| COVID-19 |  |  |  |
| COVID-19 (All cases vs population) | 0.93 (0.89-0.98) | 0.009 | 159,840 / 2,782,977 |
| COVID-19 (Hospitalised) | 0.83 (0.74-0.93) | 0.002 | 44,986 / 2,356,386 |
| COVID-19 (Severe respiratory vs population) | 0.69 (0.57-0.84) | 0.000 | 18,152 / 1,145,546 |
| Infection |  |  |  |
| URTI | 1.67 (1.13-2.48) | 0.010 | 2,795 / 483,689 |
| Endocarditis | 1.66 (0.88-3.15) | 0.120 | 1,080 / 485,404 |
| Cholecystitis | 1.53 (1.12-2.08) | 0.007 | 4,052 / 482,432 |
| UTI | 1.32 (1.17-1.5) | 0.000 | 21,958 / 464,256 |
| Osteomyelitis | 1.27 (0.88-1.82) | 0.207 | 4,836 / 481,648 |
| Appendicitis | 1.21 (0.93-1.58) | 0.153 | 4,604 / 481,880 |
| Cellulitis | 1.2 (1.02-1.41) | 0.028 | 12,196 / 474,288 |
| LRTI | 1 (0.83-1.22) | 0.960 | 14,135 / 472,349 |
| Pneumonia | 0.97 (0.86-1.1) | 0.642 | 22,567 / 463,917 |
| Sepsis | 0.8 (0.66-0.96) | 0.019 | 11,643 / 474,841 |
| Other critical care outcomes |  |  |  |
| Pneumonia (critical care) | 0.69 (0.49-0.97) | 0.034 | 2,758 / 428,607 |
| LRTI (critical care) | 0.51 (0.23-1.12) | 0.095 | 585 / 430,780 |
| Pneumonia (28-day death in critical care) | 0.5 (0.23-1.08) | 0.077 | 803 / 430,820 |

After weighting, we performed Two Sample Mendelian randomisation analyses using this instrument on each outcome in turn, comparing results to that generated using the *cis*IL6R instrument.

### Sensitivity analyses

We performed three broad types of sensitivity analysis. Firstly, we attempted to test the assumptions of MR using alternative meta-analytic approaches (MR-Egger and weighted median approaches).

Secondly, we tested whether variant weighting altered the results, by weighting variants solely on their effect estimates from CHARGE rather than from the UKB-CHARGE meta-analysis to avoid over-fitting of data (“winners curse”),^21^ and c) ran the analysis weighted entirely on the SNP-outcome association (e.g. unweighted by CRP), Finally, we tested whether our choice of SNPs to include altered the results. Firstly, we used the R package *RadialMR* to identify SNP outliers, and re-ran analyses with outliers excluded.^37^ Secondly, we performed iterative leave-one-out analyses, where each SNP is left out of the model in turn, and IVW estimates recalculated. Thirdly, we re-ran analysis re-ran the analysis including only variants within 10kb of *IL6R*, and finally, we ran the canonical and well described Asp358Ala SNP (rs2228145) as a single instrument.^16^

### Data availability

Most data used in this study are publicly available. For ease, curated data (harmonised SNPs and MR results) are available at the authors GitHub (https://github.com/gushamilton/il6-sepsis), so all findings can be replicated. Raw data from the FinnGen GWAS are available via the FinnGen website (https://www.finngen.fi/), COVID-HGI GWAS are available via the COVID-HGI website (https://www.covid19hg.org/), and the UK Biobank GWAS performed as part of this study are available at the MRC-IEU Open GWAS repository (https://gwas.mrcieu.ac.uk/). Access to the full summary statistics for the sepsis GWAS performed by the GaINS and GenOSept committee is by application to the relevant committee. This research was performed under UK Biobank application 56243. Individual access to UK Biobank can be arranged via the UK Biobank website.

### Reporting guidelines

This study is reported in line with the STROBE-MR guidance, with the checklist available in the supplement.^38^

## Results

### Identification of sepsis cases in UK Biobank

In UK Biobank, we identified 11,643 cases of sepsis, with 474,841 controls of European ancestry. 1,896 patients died within 30 days of admission, with 484,588 controls. 1,380 patients had critical care admission with sepsis, with 429,925 controls (including only UK Biobank participants in England). 347 patients died within 30 days of critical care admission, leaving 431,018 controls. We subsequently performed a case-control GWAS for each of these outcomes, with links to quantile-quantile plots and Manhattan plots available in **Supplementary Table 2**, as well as details of other included infections in **Table 1**.

### Mendelian randomisation

We performed inverse variance weighted (IVW) Mendelian randomisation on each outcome in turn. As our instruments are weighted by high-sensitivity CRP (hsCRP), odds ratios (OR) are on the scale of natural log hsCRP decrease. For reference, we call this IL6R blockade, with odds ratios of more than one representing increasing risk with greater interference and odds ratios of less than one representing decreasing risk.

For our primary outcome - sepsis - we identified a severity dependent effect, with evidence suggesting that IL6R blockade is increasingly protective with more severe disease. The Odds Ratio (OR) for sepsis was 0.80 (95% CI 0.66-0.96); for 28-day death in sepsis was 0.74 (95% CI 0.47-1.15), for sepsis requiring critical care admission was 0.48 (95% CI 0.38-0.78) and for 28-day death in sepsis requiring critical care admission was 0.37 (95% CI 0.14-0.98) (Figure 2A, Table 1).

**Figure 2:**
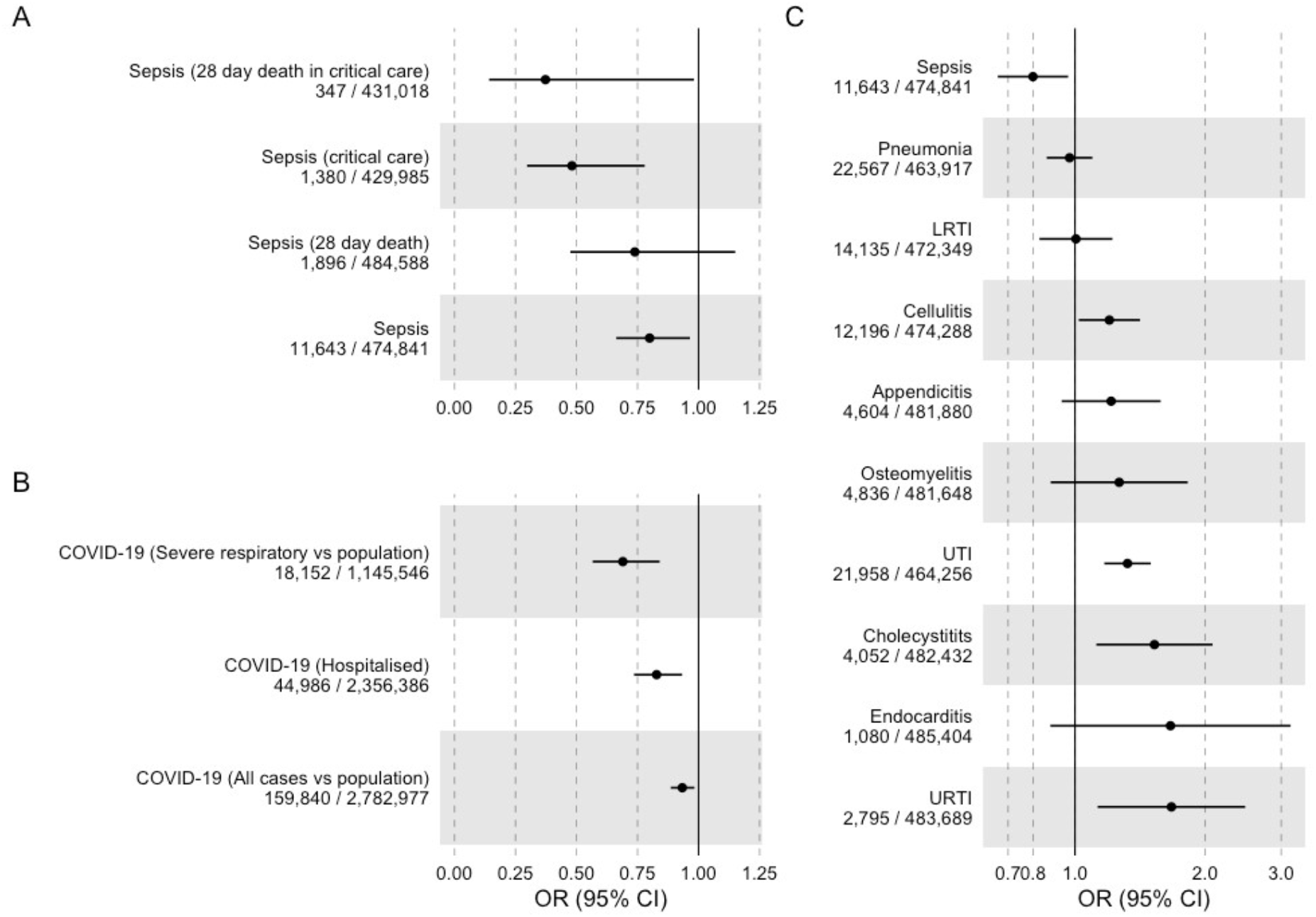
IVW MR estimates (Odds Ratios; ORs) with 95% confidence intervals (CIs) for IL6R blockade and each outcome (A: Sepsis, B: COVID, C: other infections) in UK Biobank

These severity dependent effects were mirrored when we performed IVW MR on COVID-19 related outcomes from the COVID-HGI, with evidence suggesting that IL6R blockade is more protective from critical respiratory illness, defined as those who required respiratory support or who died during hospitalisation (OR 0.69, 95% CI 0.57-0.84) than from hospitalisation alone (OR 0.83, 95% CI 0.74-0.93). Notwithstanding uncertainty around the point estimates, MR estimates for IL6R blockade were larger in sepsis than in COVID-19, for the comparable disease severity.

We performed IVW MR to investigate the association of IL6R blockade on the odds of infection in the absence of sepsis within UK Biobank. In line with trial and registry literature, there was evidence suggesting an increase in susceptibility to evaluated infections. The largest effect sizes observed were for URTI (OR 1.67; 95% CI 1.13-2.48) and for endocarditis (OR 1.66; 95% CI 0.88-3.15). For respiratory infections – LRTI and pneumonia - we saw no strong evidence of effect (OR ∼ 1 for both) (Figure 2C, Table 1).

For both of these respiratory infections, despite the largely null effect with incidence of disease, IL6R blockade was associated with decreased odds of critical care admission (Figure 3A, Table 1), with effect estimates concordant with the estimates from sepsis requiring critical care admission: OR for pneumonia requiring critical care admission was 0.69 (95% CI 0.49-0.97) and the OR for LRTI requiring critical care admission was 0.51 (95% CI 0.23-1.21).

**Figure 3:**
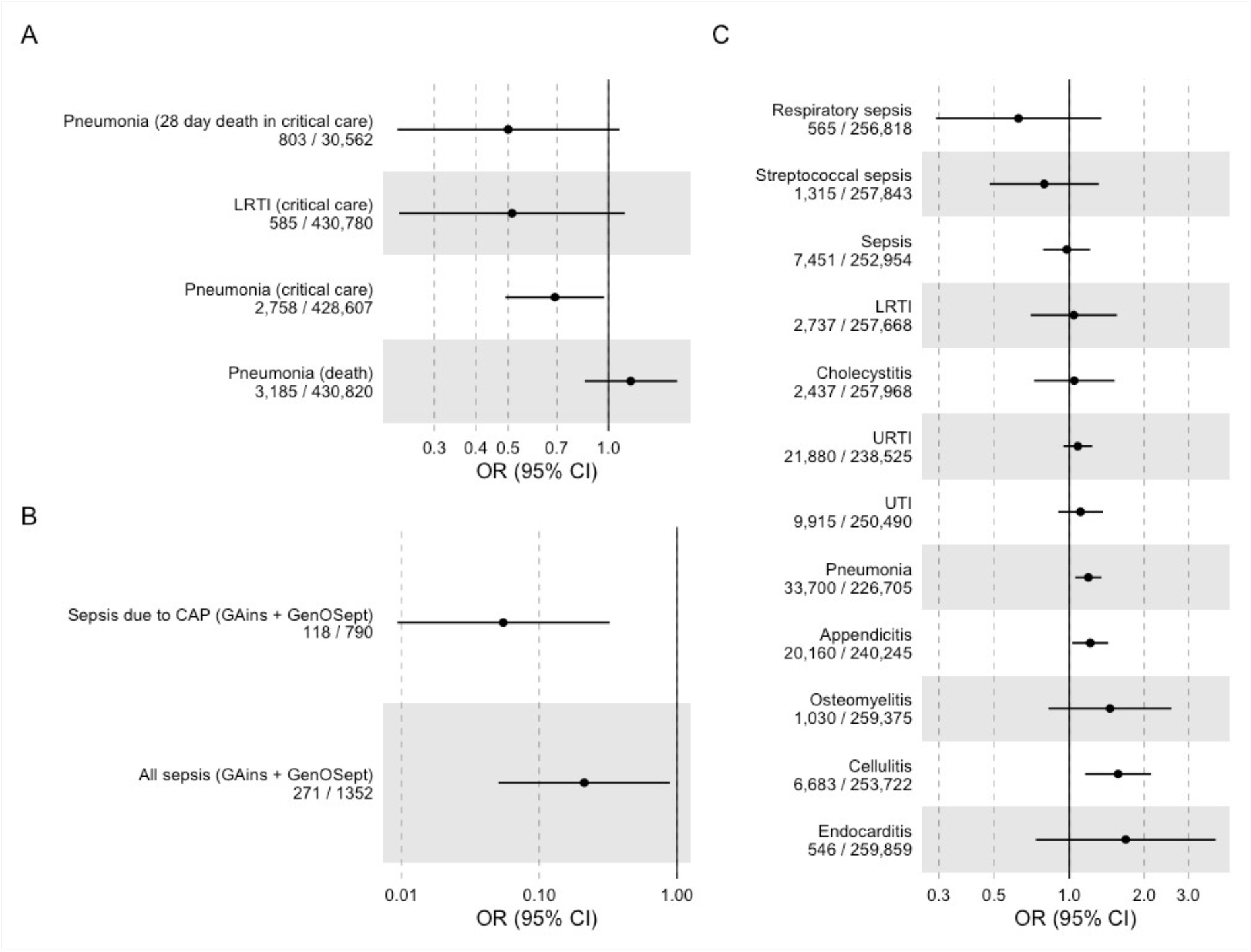
Inverse variance weighted MR estimates of IL6R blockade with 95% confidence intervals (CIs) for a) respiratory infection, b) survival from sepsis related to critical care admission, and c) FinnGen replication cohort

### Follow-up analysis

We were able to extract parallel GWAS results from Round 6 of the FinnGen consortium (**Supplementary Table 3**) and to re-run all analyses. The frequency of incident cases differed across infection between studies. This was most notable for LRTI (frequency of incident cases 2.9% UK Biobank compared to 1.05% FinnGen). We identified three sepsis outcomes to compare: one combined one (all cases of ICD-coded sepsis) and two specific ones (subsets of the combined outcome). For the combined sepsis outcome the frequency of incident cases was 54% higher in FinnGen (2.4 % UK Biobank, 3.7% FinnGen), while the frequency of mortality at 5 years - the only mortality figure available within FinnGen – was 20.2% as opposed to 37.6% within UK Biobank.

For the specific sepsis definitions available in FinnGen - sepsis due to pneumonia and streptococcal sepsis, mortality data was not available, but these were rarer than the combined sepsis outcome (0.5% and 0.2% respectively).

For the combined FinnGen sepsis outcome, the odds ratio was close the null: OR 0.98 (95% CI 0.79 - 1.21). However, when focusing on the two specific outcomes of Streptococcal sepsis and sepsis due to pneumonia, effect estimates were similar to those using UK Biobank: OR 0.79 (95% CI 0.48 - 1.31) and OR 0.63 (95% CI 0.29-1.35) respectively although with more uncertainty due to a smaller sample size. (**Figure 3C**).

Across UK Biobank and FinnGen, effect estimates were largely consistent in direction for all infections except pneumonia although effect estimates were generally smaller. The meta-analysed summary estimate for sepsis, utilising UK Biobank and the main FinnGen sepsis outcome had an OR of 0.86 (0.74-0.99), when meta-analysing with streptococcal sepsis in FinnGen the OR was 0.80 (0.67-0.95) and when meta-analysing with respiratory sepsis in FinnGen the OR was 0.79 (0.66-0.95). FinnGen specific and meta-analysed summary effects are available in **Table S4**, with a summary forest plot in **Figure S1**.

### Additional survival outcomes

Additional survival outcomes were available from two previous GWAS performed in the GAiNS and GenOSept consortium, both of which recruited patients with sepsis in critical care, but also included a subset of patients with confirmed CAP. In both studies, case-control GWAS was performed with the outcome of 28-day survival.

In both studies, we performed IVW MR and effects were concordant with our primary data (Figure 3B), with a summary OR of 0.22 (95% CI 0.04-1.31) for sepsis and 0.06 (95% CI 0.010.55) for the CAP subset. These estimates were imprecise, given the number of included cases and controls.

### Alternative genetic instruments

Although our genetic instrument includes variants around *IL6R* (acting as a proxy for the target drug effect), we weighted it by the effect on hsCRP as this represents an appropriate read-out of IL6R function and the effect on downstream IL-6 signalling. However, this does not mean that hsCRP is itself necessarily part of the causal pathway and it may simply represent a measurable marker of IL6R blockade. It is also plausible that the beneficial effect of IL-6 blockade in sepsis is actually mediated by reductions in hsCRP. In an attempt to interrogate this potential mediation and elucidate a potential mechanism of effect of IL6R blockade we re-ran analyses using established cis variants around *CRP* known to be associated with circulating hsCRP levels.

In IVW MR analyses using the *cis*CRP instrument, we found evidence that CRP may be a part of the IL6R blockade effect, with evidence of reduced odds of sepsis and sepsis related mortality: OR for sepsis outcome 0.91, 95% CI 0.82 - 1), OR for sepsis critical care admission 0.88, 95% CI 0.65-1.17), OR for death 0.72, 0.59 - 0.93), OR for critical care death 0.58, 95% CI 0.32 – 1.04 (**Figure 4, Supplementary Table 6**). This effect was also seen in FinnGen: OR for combined sepsis outcome 0.76, 95% CI 0.59-0.98), with similar results for other sepsis definitions. These effects were generally smaller in size than those seen using the *cis*IL6R instrument, with less evidence of a graduated effect with severity as seen with the *cis*IL6R instrument. Effect estimates were again broadly similar (although imprecise) for protection against critical infection (e.g. OR for critical care admission with LRTI 0.77, 95% CI 0.49-1.21).

**Figure 4:**
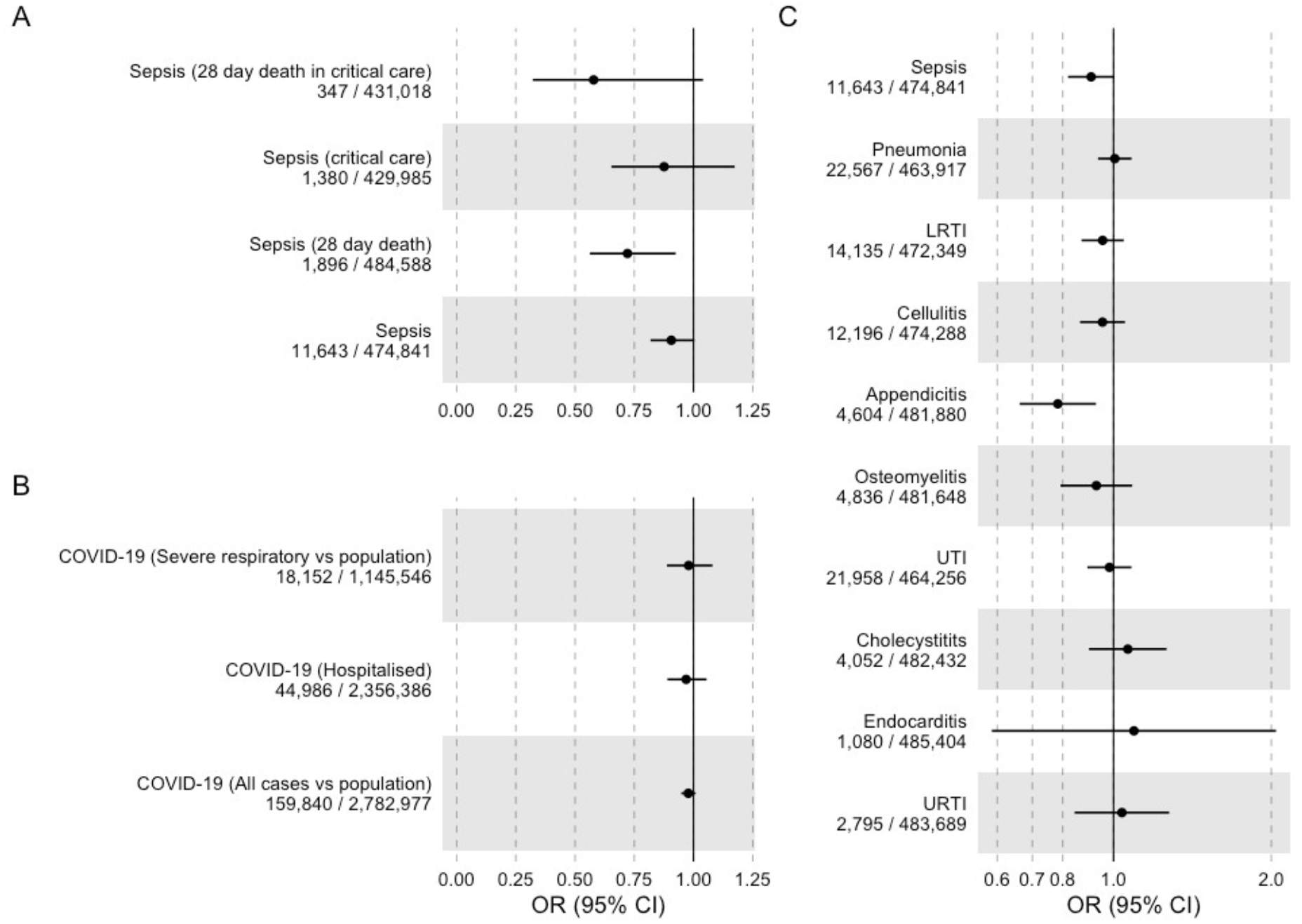
Inverse variance weighted MR effect estimates for CRP from *cis*CRP variants and each outcome (A: Sepsis, B: COVID, C: other infections)

Outside of sepsis, there was little evidence of correlation between IVW MR results from *cis*CRP and *cis*IL6R related instruments (Pearson’s R 0.06), and in addition, there was no strong evidence for an association with COVID-19 outcomes.

### Sensitivity analyses

We performed a range of sensitivity analyses which did not alter the interpretation of primary results. Firstly, where possible, we tested the assumptions of Mendelian randomisation using different meta-analytic techniques.^21^ For these analyses, we performed meta-analysis using MR Egger and Weighted Median approaches. Weighted median analyses were broadly similar to IVW results, but MR-Egger estimates were more imprecise, with nearly all confidence intervals for outcomes in UK Biobank, FinnGen, and COVID-HGI crossing the null. These results are shown in **Supplementary Figure 2** and **Supplementary Table 7**, while representative scatter plots for two representative outcomes (sepsis and critical care admission with sepsis) are available in **Supplementary Figure 3**.

In a second set of sensitivity analyses, we tested whether the way we designed and weighted our instrument materially affected the results. Firstly, we weighted our exposure SNPs using betas solely from the CHARGE consortium^34^ (rather than using a combined CHARGE-UK Biobank meta-analysis, **Supplementary Table 8**). These results were similar (although more imprecise, due to the smaller sample size of the original GWAS). Secondly, we ran completely unweighted analyses (**Supplementary Table 9, Supplementary Figure 4**), which simply meta-analysed the effect of each SNP-outcome association, unweighted by any downstream effect. These results were again concordant with our primary analysis, although were highly imprecise (given the small individual effect of each SNP).

In a third set of sensitivity analyses, we tested whether our selection of *IL6R* SNPs influenced results. Firstly, we identified and removed outliers using Radial MR, and re-ran analyses (**Supplementary Figure 5, Supplementary Table 10**). We identified no alteration in our primary results. Alongside that, we performed leave one out analyses, removing individual SNPs from the analyses and re-running IVW MR. For ease, we show this for the first four outcomes only in **Supplementary Figure 6**, which show the removal of any individual SNP does not alter the main results. Finally, we restricted analyses to the seven SNPs within 10kb of *IL6R*, (**Supplementary Table 11**) and the rs2228145 SNP, which has a known functional effect on IL6R and downstream effects^36^ (**Supplementary Figure 7, Supplementary Table 12**). Again, these effects were all concordant with our primary results, although with reduced power as expected by removing multiple SNPs from the meta-analysis.

## Discussion

In this study, we provide evidence from multiple, independent data sources, that blockade of IL-6 signalling pathways is likely to be protective against the development of sepsis. There is evidence across both UK Biobank and FinnGen that the apparent protective effect of functional IL6R blockade increases with increasing severity of illness with potential protective effects on short term death in sepsis, and critical care admission with sepsis. This effect was similar (although slightly larger) than estimates relating to severe COVID-19, where IL6RA have already been shown to improve mortality.^11^ In contrast, functional IL6R blockade appears to show evidence suggesting increases in susceptibility to infections, matching trial and registry data. We also found evidence that IL6R blockade may be protective in critical illness in respiratory infection, where effect estimates were similar to those seen in sepsis and consistent with the COVID-19 data. Given the similarities between COVID-19 pneumonitis, sepsis, and bacterial respiratory infection and with commonalities in underlying pathophysiology, this suggests IL6RA as a potentially broad therapeutic target for patients unwell with critical infection.

Our analysis using *cis*CRP instruments had similar findings, although effect estimates were generally smaller. This suggests that one potential route by which IL6R blockade reduces the odds of severe sepsis is by reducing CRP, although this remains a hypothesis. Given the ongoing development of therapeutics that target trans-specific IL-6 signalling, and therapeutics that target CRP itself, this may represent a future avenue in sepsis therapeutics^39,40^, however this does not alter the interpretation of the primary IL6R blockade related finding here. Additionally, the concordance of MR effect estimates between *cis*CRP and *cis*IL6R genetic variants provides confidence in our primary analysis, and strongly supports the role of IL-6 signalling in sepsis.

Previous literature has not identified an association between variants proxying IL6R blockade and sepsis, although multiple studies have identified associations (in line with our estimates), suggesting these *IL6R* variants increases the risk of infection, which match randomised trial data.^30,41^. One trial was considering the use of IL6RA in paediatric sepsis (NCT04850443), but this was halted due to lack of funding. Furthermore, randomised trials have been performed aiming to remove IL-6 and other cytokines by using extracorporeal haemoadsorption devices.^42^ In the largest trial (97 evaluable patients), the use of haemoadsorption in patients with severe sepsis was not associated with any reduction in plasma IL-6 levels and had no effect on mortality once adjusting for comorbidities (Hazard Ratio 1.67, 95% CI 0.77 - 3.61).^43^ Given that this device is untargeted, did not successfully reduce IL-6 levels and the clinically relevant complications associated with usage of extracorporeal haemoadsorption, it is hard to interpret this evidence in evaluation of targeted IL-6 downregulation by either genetic variation or IL6RA.

Despite a large sample size, and multiple independent sources of data, this study has weaknesses. Firstly, we rely on diagnostic coding for infections, which varies between and across cohorts. Importantly, across most infections, estimates were broadly similar. In sepsis, our estimates in our replication cohort were dependent on the definition of sepsis, with much larger effects in those with specific sepsis codes (e.g. streptococcal sepsis) than with the main code. However, this smaller effect may be due to the FinnGen sepsis cases being of lower severity than the UK Biobank cohort, reflected in the higher frequency of incident cases of sepsis, and reduced severity of disease. Given our results show IL-6 activity has both protective (from infection) and detrimental (development of sepsis) effects, even small changes to include less sick populations will likely greatly reduce the effect size, as seen in the FinnGen data and demonstrated in studies and simulations of phenotypic misclassification.

These results are subject to other factors which potentially complicate the translation of applied epidemiological analysis into clinical trials. This is particularly acute with respect to the potential definition of a population that might benefit from IL-6 inhibition in the context of the severity of sepsis, but also the possible risks according to other infection risk (especially in the frail). This remains a question not addressed directly by the work here, but of potentially great importance if IL6RA are to be considered as interventions for acute outcomes.

Related to this, a major challenge also exists in interpreting potential biases induced with the analysis of sepsis which is a product of case status – i.e. of severe infection. As the downregulation of functional IL6R potentially leads to increased risk of infection, analyses of genetic variants related to progression to severe infections (e.g. sepsis) has the potential to be biased. In extreme cases, collider bias could lead to unpredictable biases on effect estimates, if the genetic effects on incidence of sepsis were very large. For that reason, we undertook GWAS using the entire cohorts, rather than performing a “case-only” analysis. This potentially avoids the impact of this specific problem, but leaves interpretation subject to results based on control status including mild infection. As a consequence, this may mean that the size effect estimates here (e.g. for apparent reduced odds of severe sepsis) should not be interpreted as the same as those potentially occurring with the use of IL6RA therapy in real life. Despite this, it is reassuring that our effect estimates are similar in size and direction to COVID-19 effect estimates, where a large and clinically important effect was identified (3-4% absolute reduction in mortality for those hospitalised with COVID-19).^11^ In the absence of a trial, these types of interpretation complication remain difficult to escape.

Focusing on variants within or near the *IL6R* it is difficult to completely rule out pleiotropic effects (e.g. these SNPs acting to reduce risk of sepsis by another, unrelated mechanism). However, given the concordance in protection from severe COVID-19 identified in our study, randomised trial data matching that using genetic proxies for IL6R blockade, and the location of these SNPs we can have some confidence that the effect is driven by alterations in IL6R.

Finally, in common to all genetic studies, translation of (small) effects due to germline variation into the context of a severely ill population receiving a large, time limited intervention, requires detailed thought. Not only does a focus on variants within or near the *IL6R* fail to guarantee the absence of complications generated as a result of pleiotropy, most of our estimates relate to protection from the odds of sepsis events. Any potential trial is likely to enrol patients who already have sepsis and given the difficulties in predicting sepsis and the practicalities of administering IL6RA, differences between those circumstances and results here may appear. Furthermore, in clinical trials of COVID-19 outcomes, participants also received corticosteroids with some evidence of an interaction effect between corticosteroids and IL6RA.^11^ Although the risk of nosocomial infection was not high in clinical trials in COVID-19 (<1% in both REMAP-CAP and RECOVERY^9,10^), careful evaluation of the potential harms of IL6RA in a population with infection will be required in trial design, given the double edged nature of IL6 inhibition.

## Conclusion

Although we should be appropriately sceptical of any novel therapeutics in sepsis, given the failure to identify any successful agents despite 30 years of research, the unique conditions surrounding this work suggest that IL6R blockade may be a useful approach. Firstly, the use of MR to identify potential therapeutic targets is supported by a large amount of empirical evidence, replicating both positive and negative trial effects.^20,44^ Secondly, our specific technique of using *IL6R* variants as an exposure has been utilised before, with randomised trial data matching MR estimates.^12,16^ Finally, the similarities between COVID-19 and sepsis pathophysiology are clear, with robust trial data supporting the role of IL6RA in COVID-19.^11^ Against expectations, adjusted rates of secondary infection in the IL6RA treated population were similar (OR: 0.99, 95% CI: 0.85, 1.16) than those in comparator arms in the recent WHO meta-analysis, which provides additional comfort that IL6RA are safe to use in the critically ill.^11^

Our data are therefore suggestive that functional downregulation of IL6 may have beneficial effects in improving sepsis outcomes. Given the previous data linking genetic variation in *IL6R* with trial outcomes of IL6RA, and the biological plausibility of effect, these data support trialling IL6RA in sepsis.

## Supporting information

STROBE statement

## Data Availability

Data availability
Most data used in this study are publicly available. For ease curated data (harmonised SNPs and MR results) are available at the authors GitHub (https://github.com/gushamilton/il6-sepsis) so all findings can be replicated. Raw data from the FinnGen GWAS are available via the FinnGen website (https://www.finngen.fi/) COVID-HGI GWAS are available via the COVID-HGI website (https://www.covid19hg.org/) and the UK Biobank GWAS performed as part of this study are available at the MRC-IEU Open GWAS repository (https://gwas.mrcieu.ac.uk/). Access to the full summary statistics for the sepsis GWAS performed by the GaINS and GenOSept committee is by application to the relevant committee. This research was performed under UK Biobank application 56243. Individual access to UK Biobank can be arranged via the UK Biobank website.

## Ethics

Most of the data in this study was publicly available and non-identifiable. Therefore, no ethical approval was required to access it. Access to UK Biobank was arranged by the UK Biobank IDAC. This study was performed under application number 56243.

