## Supplementary Figures for "Therapeutic potential of IL6R blockade for the treatment of sepsis and sepsis-related death: Findings from a Mendelian randomisation study"

Figure S1: Meta-analysed effects of across UKB and FinnGen

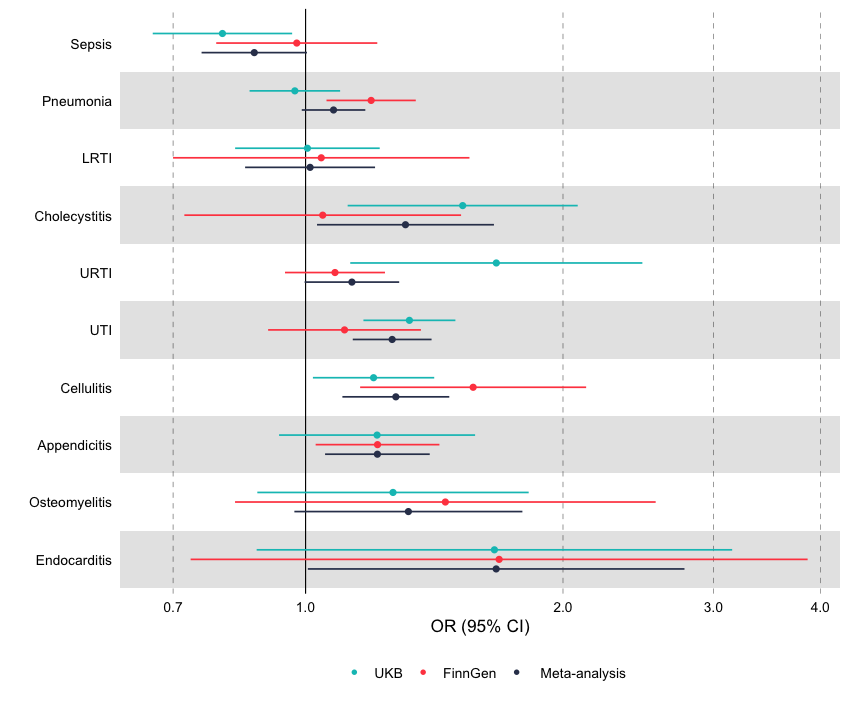

Figure S2: Weighted median and MR Egger effect estimates for the UK Biobank, COVID-19 and FinnGen outcomes.

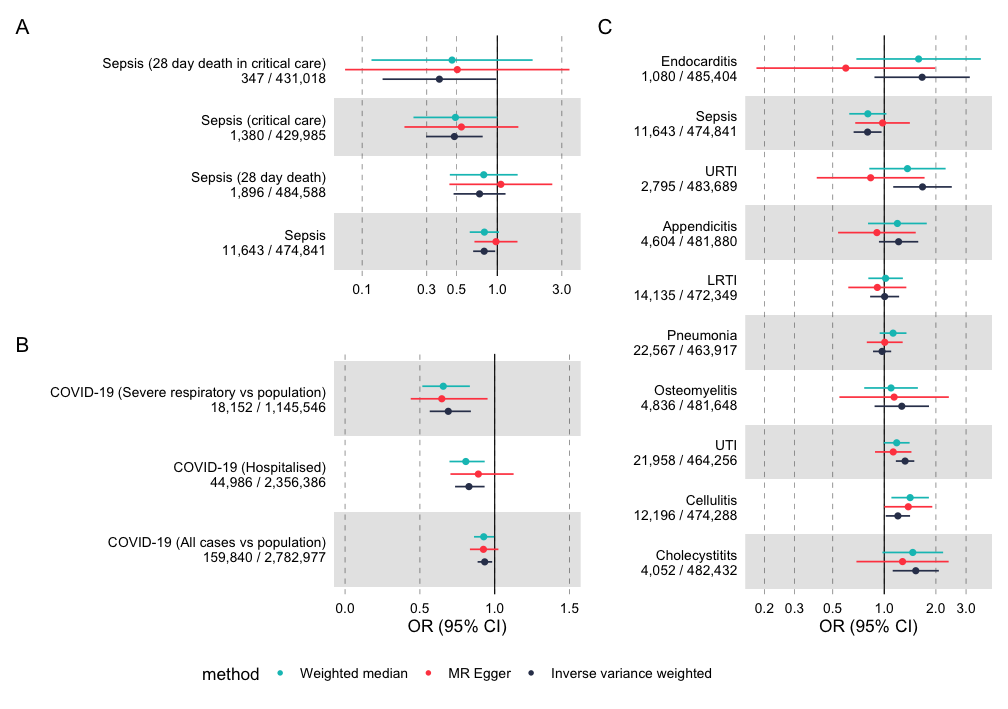

Figure S3: MR Scatter plots for sepsis and critical care admission with sepsis
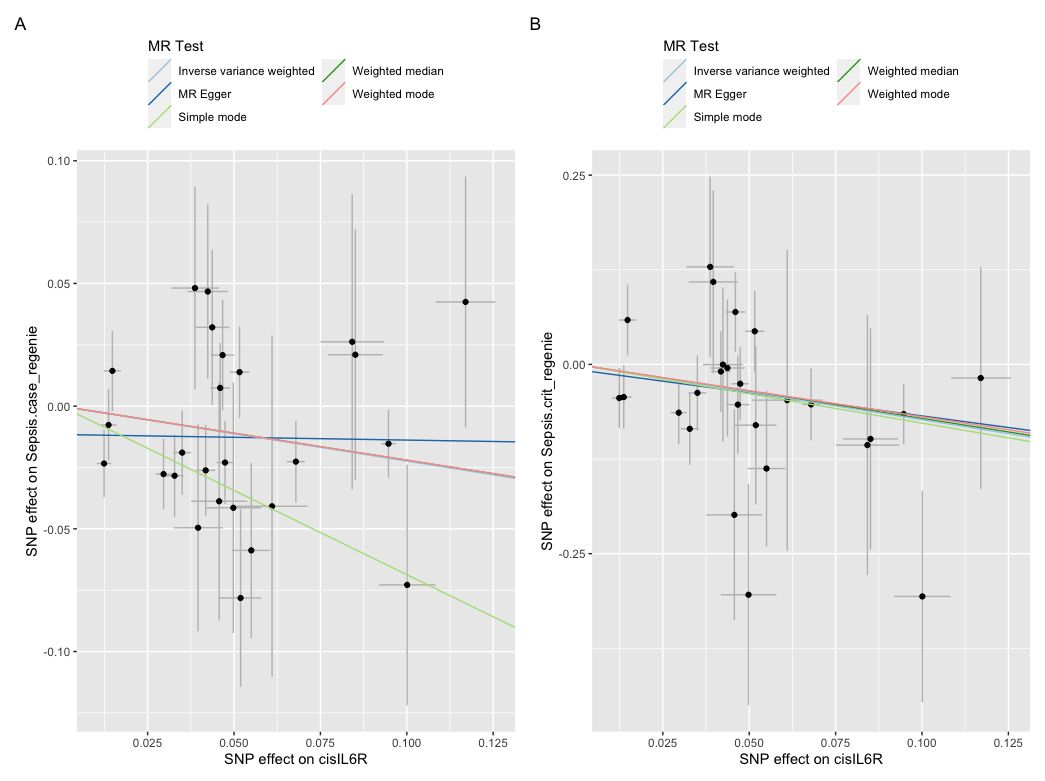

Figure S4: Unweighted inverse variance weighted estimates of cisIL6R variants and effect on the primary outcome

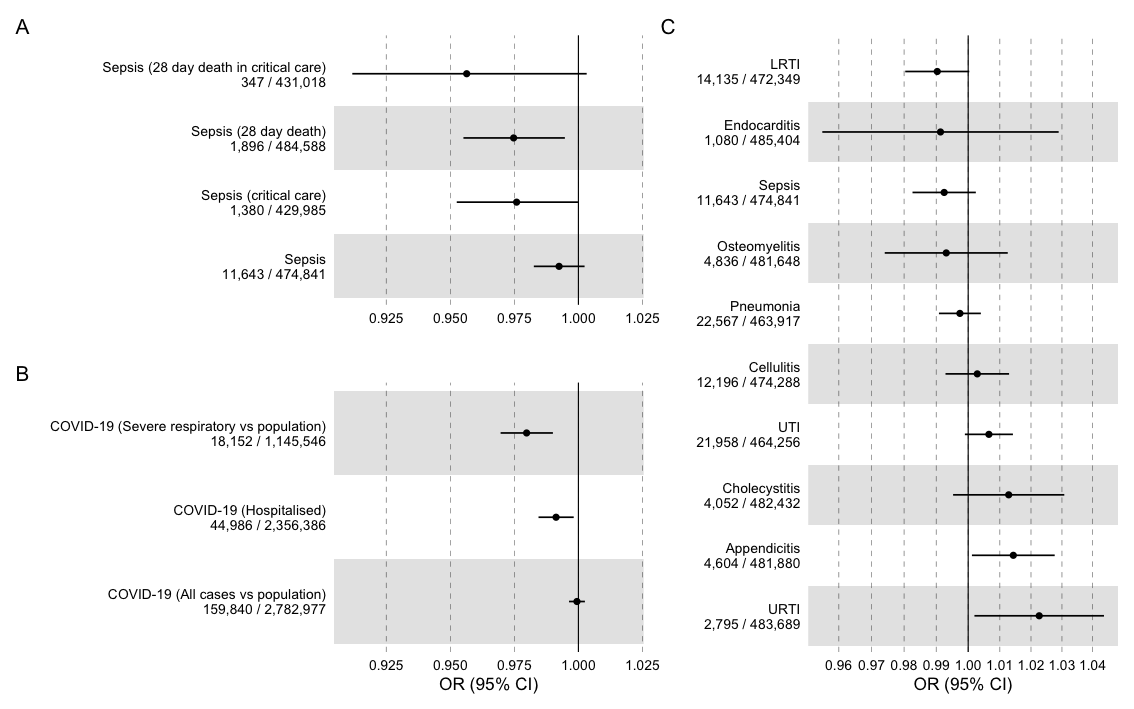

Figure S5: Plots removing outliers detected by RadialMR

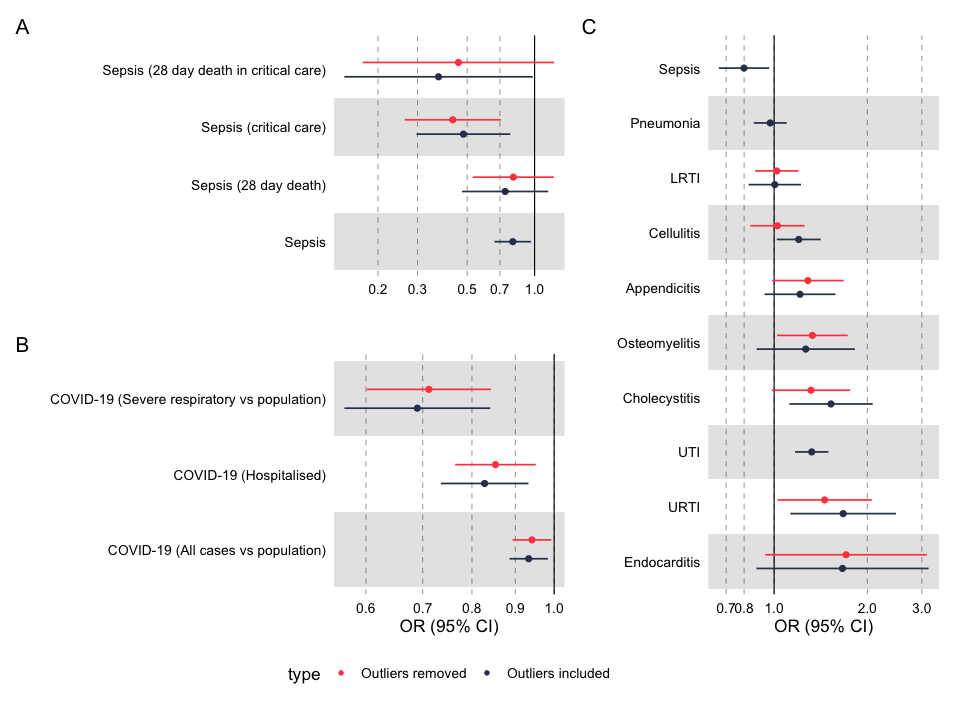

Figure S6: Leave one out analyses for a: critically unwell sepsis cases, b: level 3 sepsis cases, c: sepsis cases, and d: sepsis related mortality.

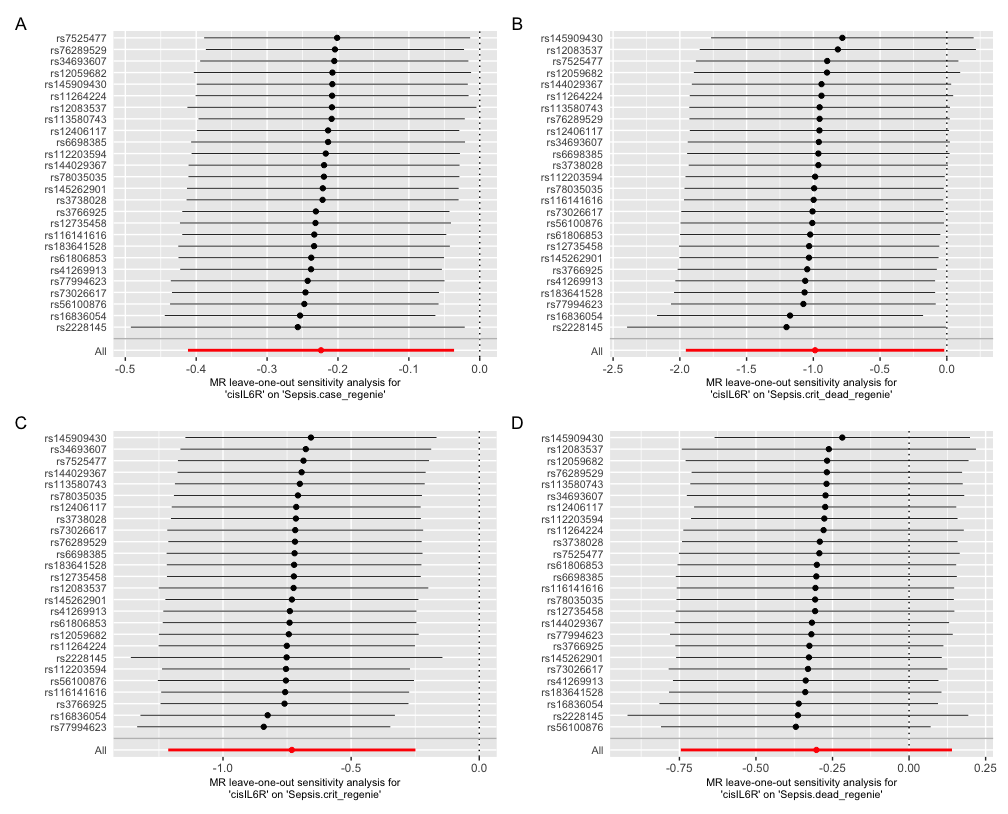

Figure S7: MR Wald ratio estimates for the Asp358Ala (rs2228145) SNP association with UK Biobank, COVID-19, and FinnGen outcomes

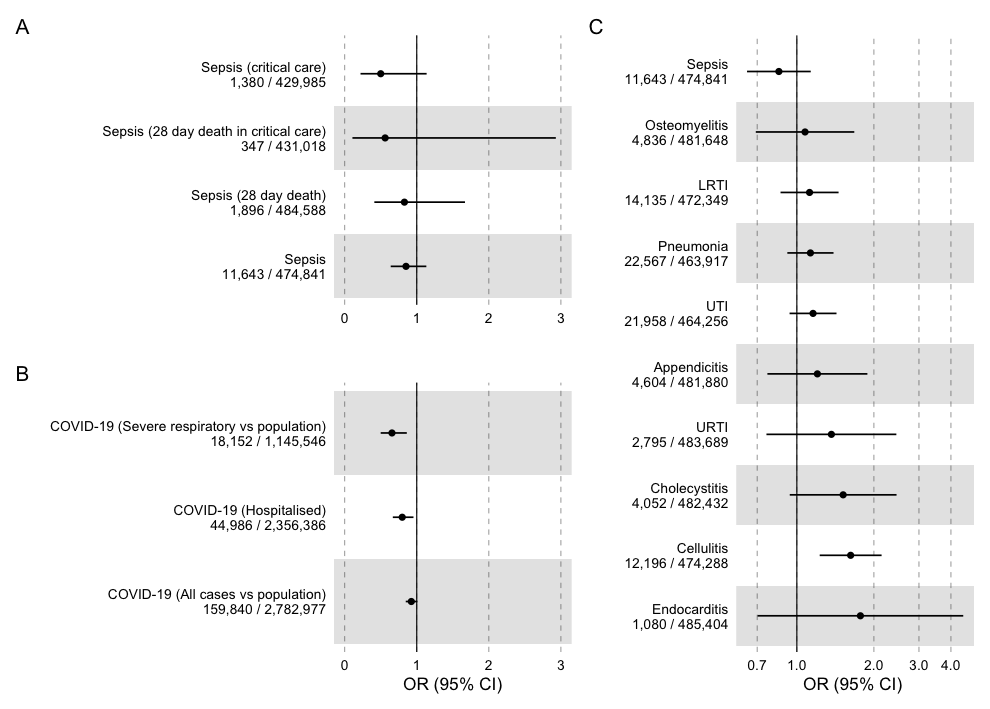
