## Supplementary Methods for "Therapeutic potential of IL6R blockade for the treatment of sepsis and sepsis-related death: Findings from a Mendelian randomisation study"

### GWAS:

### Genotyping and imputation

The data release for this analysis contains the cohort of successfully genotyped samples (n=488,377). 49,979 individuals were genotyped using the UK BiLEVE array and 438,398 using the UK Biobank axiom array. Pre-imputation QC, phasing and imputation are described elsewhere.^23^ In brief, prior to phasing, multiallelic SNPs or those with MAF ≤1% were removed. Phasing of genotype data was performed using a modified version of the SHAPEIT2 algorithm. Genotype imputation to a reference set combining the UK10K haplotype and HRC reference panels was performed using IMPUTE2 algorithms.

The analyses presented here were restricted to autosomal variants using a graded filtering with varying imputation quality for different allele frequency ranges. Therefore, rarer genetic variants are required to have a higher imputation INFO score (Info>0.3 for MAF >3%; Info>0.6 for MAF 1-3%; Info>0.8 for MAF 0.5-1%; Info>0.9 for MAF 0.1-0.5%) with MAF and Info scores having been recalculated on an in-house derived ‘European’ subset.^23^

### Data quality control

Individuals with sex-mismatch (derived by comparing genetic sex and reported sex) or individuals with sex-chromosome aneuploidy were excluded from the analysis (n=814).

### Ancestry

We restricted the sample to individuals of ‘European’ ancestry as defined by an in-house kmeans cluster analysis performed using the first 4 principal components provided by UK Biobank in the statistical software environment R. The current analysis includes the largest cluster from this analysis (n=464,708).^23^

### GWAS methodology

regenie was chosen to perform GWAS as it reduces type 1 error when there is significant case control imbalance, as compared to linear mixed model approaches, with prior data suggesting good performance with case-control ratios as high as 1:660 in UK Biobank, as well as controlling for relatedness.^24^

GWAS was performed using regenie 2.2.2 adjusting for age, sex, genetic chip, UK Biobank assessment centre and the first ten principal components, with further details in the supplement. LD Score regression (v1.0.1) was then used to calculate genomic inflation.^25^

*Definition and justification of instrument*

We chose to weight this downregulation by the effect of each variant on hsCRP. The weighting on hsCRP was performed for multiple reasons. Firstly, IL6 is responsible for the production of hsCRP in hepatocytes and hsCRP is directly downstream of IL6, and hsCRP levels therefore reflect the activity of the IL6 pathway.^19,44,45^ Secondly, trial data shows large effects of IL6RAs on circulating hsCRP in a wide range of clinical settings, with hsCRP levels often correlating with clinical outcomes.^12,13,46,47^. Finally, this approach (*IL6R* variants weighted by CRP to proxy IL6RA therapy) has been recommend and/or utilised in multiple recent studies, and there are large-scale GWAS available to provide precise estimates.^18–^

20,30,33,44,45

This weighting on CRP therefore does not make an assumption of any causal pathway, however, reflects a best-available handle on the functional effect of each IL6R variant on IL6 downregulation and hence the appropriate weight for inclusion in follow up MR analysis. In further analyses, described below, we assess whether hsCRP might be mediating the IL6R effect using *cis*CRP variants.

Conceptually our genetic instrument is similar to the action of anti-IL6R monoclonal antibodies (e.g. tocilizumab) that lead to complete inhibition of IL6R signalling by blocking both IL6 classical and trans signalling.^17–19,44,45^ For simplicity we henceforth refer to this exposure as “IL6R blockade”. It is recognised that this is an oversimplification of the IL6 pathway, with evidence that effects on health outcomes are mediated by classical and trans signalling in differing ways, and that the effect of our instrument may not act in the same way as IL6RAs.^8^ In our sensitivity analyses (below), we explore other ways of defining our exposure, including alternative weighting strategies.

*Secondary cohorts*

We included three secondary cohorts in this study: Firstly, we extracted GWAS summary statistics from R6 of the FinnGen consortium for each of the infections included in the UK Biobank analysis and for sepsis. Code lists used to define each outcome, variants included, and GWAS methodology are available at the FinnGen website^1^ and in **Supplementary Table 2**

For our COVID-19 analysis, we extracted summary statistics from the COVID Host Genetics Initiative Round 7, a worldwide consortium that included patients with COVID-19 from a variety of studies.^27^ Four separate case-control meta-analyses were run: a) very severe confirmed COVID-19 requiring respiratory support vs population; 18,152 cases vs 1,145,546 controls b) hospitalised COVID-19 vs population (44,968 cases vs 2,356,386 controls), and c) COVID-19 cases vs population (159,840 cases vs 2,782,977 controls). Details on included studies, populations, and methodology are available with the relevant publication.^27^

Finally, we included summary statistics from a recent genome wide association study on survival from sepsis, which included 2,553 patients from three cohorts, with various definitions of case and control, kindly provided by the GenOSept consortium.^29^ For their main analysis, they performed a GWAS of 28-day survival in sepsis due to pneumonia (359 cases vs 1,194 controls). As this GWAS was performed using SNPTEST2, it did not account for population structure as well as more recent GWAS approaches, leading to inflated p-values at low minimum allele frequencies. Therefore, LD score regression was performed, and estimates adjusted by the LD score regression where required, as described in Supplement S1.

For all secondary cohorts, if SNPs were not available in the outcome GWAS, SNP proxies (R^2^ > 0.8) were generated, with effect alleles harmonised using the *TwoSampleMR* package in R, v

0.5.6).

*Details on genetic approach*

*FinnGEN*

A detailed genetic QC approach is detailed here: https://finngen.gitbook.io/documentation/methods/phewas/quality-checks.

Full details of the GWAS methodology are available at the FinnGEN website (<https://finngen.gitbook.io/documentation/methods/phewas>). Briefly, SAIGE v 0.39.1, a was used correcting for age, sex, the first ten principal components, and genotyping batch.

*COVID-19 HGI*

Full details on the COVID-19 HGI analysis are detailed in their publication and at the COVID HGI website.^2^ Briefly, studies performed their own GWAS methodology (preferentially using regenie, and adjusting for relevant covariates), and these were meta-analysed to generate summary statistics.

*Sepsis survival GWAS – GenOSept and GAiNS.*

This data was kindly provided by the authors (Professor Charles Hinds) of a recent GWAS on sepsis survival in intensive care units, described here. As this represented separate cohorts, their initial analysis involved imputing variants using SNPTEST2 for both GAinS and GeNoSept together. Details on their methodology are at the relevant publication.^3^

*LDSC regression*

For this analysis, we also performed LD Score regression to adjust test statistics. As this data includes low number of participants, and our exposures included some SNPs with low MAF ~ 0.01, there was significant risk of p value inflation. QQ plots for a representative analysis (GaINS CAP subset) are shown below for each MAF.

MAF > 0.05

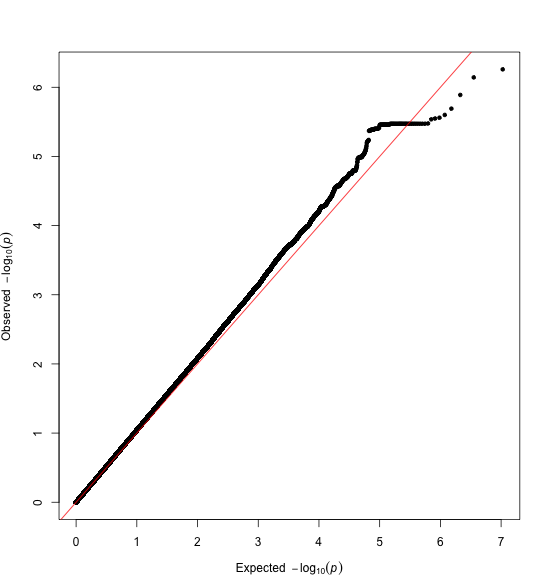

MAF < 0.05

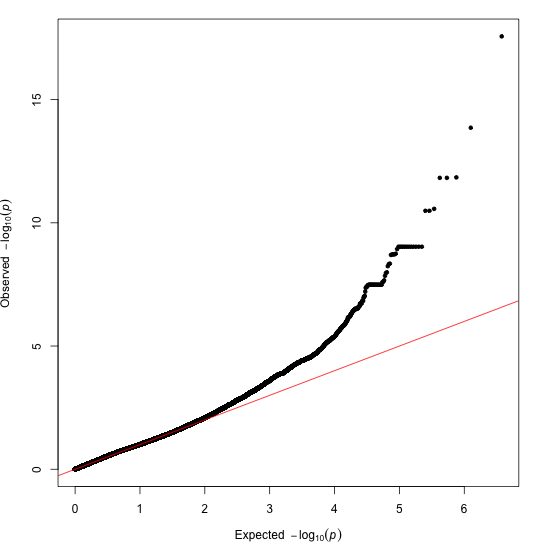

As this was likely to lead to inflated effect estimates, driven by inappropriately low p-values, we performed LD Score Regression on each dataset, using LDSC v1.0.1, and generated the LDSC intercept. LDSC intercepts for each GWAS are here:

| GAINS (CAP subset) | Intercept: 1.0391 (0.0061) |
| --- | --- |
| GAinS (All cases) | Intercept: 1.0682 (0.0063) |
| GeoSept (CAP subset) | Intercept: 1.001 (0.0069) |
| GenoSept (All cases) | Intercept: 1.005 (0.0073) |

As the GAiNS cohort had regression intercepts significantly more than 1, we adjusted each chi-sq test statistic by dividing it by the intercept and recalculated the p value. This has the effect of making the p value slightly larger.

*COVID-19 HGI*

1.	Risteys FinnGen R5 - AB1_SEPSIS [Internet]. [cited 2022 Jun 21];Available from: https://r5.risteys.finngen.fi/phenocode/AB1_SEPSIS

2.	COVID-19 Host Genetics Initiative. Mapping the human genetic architecture of COVID-19. Nature [Internet] 2021;600(7889):472–7. Available from: http://dx.doi.org/10.1038/s41586-021-03767-x

3.	Rautanen A, Mills TC, Gordon AC, et al. Genome-wide association study of survival from sepsis due to pneumonia: an observational cohort study. Lancet Respir Med [Internet] 2015;3(1):53–60. Available from: http://dx.doi.org/10.1016/S2213-2600(14)70290-5
